# Olanzapine Combined with 5-HT3 RA Plus Dexamethasone for Prevention and Treatment of Chemotherapy-Induced Nausea and Vomiting in High and Moderate Emetogenic Chemotherapy: A Systematic Review and Meta-Analysis of Randomized Controlled Trials

**DOI:** 10.1101/19009456

**Authors:** Jian-Guo Zhou, Lang Huang, Su-Han Jin, Cheng Xu, Hu Ma

## Abstract

**Objectives:** We performed a pooled analysis to evaluate the efficacy and adverse events (AEs) of olanzapine combined with dexamethasone plus compared with 5-HT3 RA plus dexamethasone for the prevention and treatment of chemotherapy-induced nausea and vomiting (CINV) in high and moderate emetogenic chemotherapy based on randomized controlled trials (RCTs).

**Methods:** PubMed, EMBASE, Web of Science, the Cochrane Library, China Biomedical Literature database (CBM), WanFang Database, China National Knowledge Infrastructure (CNKI), and Chinese Science and Technology Periodical Database (VIP) (from their inception to April 2019) were searched to capture relevant articles. Relative risk (RR) with 95% confidence intervals (CIs) for CINV and AEs were all extracted or calculated.

**Result:** Eleven studies with 1107 cancer patients were involved in this review. The pooled RR of delayed CINV (RR=0.50, 95%CI: 0.38-0.66, *P*<0.01) were significant decreased in olanzapine group. Besides, the occurrence of insomnia was statistically decreased. The rate of acute CINV (RR=0.60, 95%CI: 0.48–0.75, *P*<0.01) was also significant decreased. However only the percent of CINV □ and CINV □ were significant deceased in acute and delayed phase. Subgroup analysis demonstrated that the efficacy was no statistically significant difference between 5mg and 10mg for olanzapine.

**Conclusion:** Olanzapine significant decreased the occurrence of CINV □ and □ and insomnia in high and moderately emetogenic chemotherapy. Compared with 10mg per day, 5mg oral may be more appropriate for patients with cancer.

## Introduction

Chemotherapy-induced nausea and vomiting (CINV) is a common adverse effect (AE) in the treatment of cancer, which can significantly impair the patient’s quality of life, resulting in poor compliance and malnutrition [1]. The incidence of acute CINV (occurrence within 24 hours of administration of chemotherapy) was 36%, and 59% for delayed CINV (two to five days after the administration of chemotherapy)[2]. 5-hydroxytryptamine type 3 (5-HT3) RA[3], dexamethasone and neurokinin-1 (NK-1) RA[4]has significantly decreased the incidence of CINV, while there is still 54.7% of patients experience nausea when 5-HT3 RA, dexamethasone and NK-1 RA are used[5].

Food and Drug Administration (FDA) approved olanzapine as an antipsychotic agent that blocks α1 adrenergic receptors, serotonin 5-HT2a, 5-HT2c, 5-HT3, 5-HT6 receptors, muscarinic receptors, dopamine D1, D2, D3 receptors, and histamine H1 receptors[5]. Blocking the receptors of dopamine D2, 5-HT2c, and 5-HT3 played the role of antiemetic, and randomized controlled trials (RCTs)[6, 7] and meta-analyses[8–10] have suggested that olanzapine is useful for patients with cancers received chemotherapy. 2016 MASCC/ESMO guidelines[11] recommend olanzapine combined with 5-HT3 RA plus dexamethasone to prevent CINV, but the level of recommendation was graded to low. NCCN Guidelines for antiemesis (Version 1.2019) also recommend olanzapine combined with a 5-HT3 RA plus dexamethasone after NK-1 RA for high and moderate emetogenic chemotherapy, however 5mg per day was consider only when 10 mg per day caused serious sedation. RCTs[12, 13] with 5mg oral a day were demonstrated effective, and randomized phase II dose-finding study[14] even find 5mg had a higher complete control in delayed CINV compared with 10mg (83.1% vs. 77.6%). Meta-analysis[15] also find 5 mg and 10 mg olanzapine exhibited similar efficacy, however three (olanzapine, 5-HT3 RA and dexamethasone) or four drugs (olanzapine, 5-HT3 RA, NK-1 RA and dexamethasone) were used, besides the level of CINV was not reported. Therefore we designed this review to evaluate the efficacy and adverse events of olanzapine combined with a 5-HT3 RA plus dexamethasone related to 5-HT3 RA plus dexamethasone for the prevention and treatment of CINV in high and moderately emetogenic chemotherapy based on currently available studies.

## Methods

This review was performed following the Cochrane Handbook for Systematic Reviews of Interventions. Moreover, we reported pooled results by the recommendations listed in the Preferred Reporting Items for Systematic Reviews and Meta-Analyses (PRISMA) statement, and Supplemental Data 1 showed the details.

### Search Strategy

PubMed, Web of Science, the Cochrane Library, EMBASE, WanFang Database, China National Knowledge Infrastructure (CNKI), Chinese Science and Technology Periodical Database (VIP) and China Biomedical Literature database (CBM) (from corresponding inception to April 2019) were searched using the following terms: “olanzapine”, “chemotherapy-induced nausea and vomiting”, “CINV”, “emesis”, “nausea”. The search strategy for PubMed was documented in Supplemental Data 2. Language or date restrictions were not imposed. The bibliographies of relevant trials and review were also browsed. Two researchers independently (LH and CX) conducted in this part, and any inconsistency was solved by consulting a third researcher (HM).

### Study Inclusion

Two reviewers (J-G Z and LH) independently screened studies based on inclusion and exclusion criteria, and any inconsistency was resolved by the third investigator (HM). Eligible studies must meet the following criteria: 1) population, cancer patients who received high and moderately emetogenic chemotherapy classified by MASCC/ESMO guidelines[11]; 2) intervention, olanzapine combined with 5-HT3 RA plus dexamethasone for the prevention and treatment of CINV; 3) comparison, dexamethasone plus 5-HT3 RA for the prevention and treatment of CINV; 4) outcome, primary endpoint: the percent of delay CINV; secondary endpoint: AEs; 5) study type, we only considered RCTs.

### Exclusion Criterion

Exclusion criteria were as follows: 1) patients with no-solid tumors; 2) other antiemetic drugs were used (ie, NK-1 RA); 3) letters to editor or meeting abstracts.

### Data Extraction

Two researchers (S-H J and CX) evaluated the eligibility of all identified studies eligible through checking abstracts and titles following to the inclusion and exclusion criteria, and full-text were read carefully for possible articles. Data involved authors, country, population, publication year, drug dosage, and duration, the percentage of acute and delayed CINV and AEs. All divergences about eligibility assessment and data extraction were solved through consulting a third senior investigator (HM).

### Assessment of Methodologic Quality

The methodologic quality was appraised using the Cochrane Collaboration’s Risk Bias assessment tool for assessing the risk of bias[16] by two researchers (J-G Z and CX), and RevMan (version 5.3) was used in this part. Seven domains (blinding of outcome assessment, blinding of participants and personnel, selective reporting, allocation concealment, random-sequence generation, incomplete outcome data, and other bias) were assessed for each study, and all of them were labeled with “high risk,” “unclear risk,” or “low risk".

### Statistical Analysis

The dichotomous outcomes were estimated using relative risk (RR) with 95% confidence intervals (CIs). *I*^*2*^ statistic and *P* value were used to estimate the level of heterogeneity of involved studies[17]. We considered heterogeneity substantial if *I*^2^ ≥50% or *P*<0.10[18]. Fixed model was used when *I*^2^ <50% or *P*>0.10, if significant heterogeneity was conducted, random model was used. On the contrary, if the noticeable difference were found in clinical characteristic and methodology, regardless of *I*^2^ statistic or *P*-value, a belief of qualitative analysis was conducted. Subgroup analysis of olanzapine dosage (5mg vs. 10mg) was also conducted. We did all analyses using R (version 3.3.0) and the “meta” package.

## Result

### Literature Research and Characteristics of Studies

As showed in supplemental data 3, a total of 787 studies of olanzapine for the prevention and treatment CINV were identified. Finally, 11 articles[12, 13, 19–27] were involved in our review according to the inclusion criterion.

All involved studies were published from 2009 to 2018, and 1107 patients with cancer were involved in this review, involving 561 patients in olanzapine olanzapine group and 546 patients in control group. Except for Mukhopadhyay et al [19], all studies were from China. All patients with solid tumors received highly or moderated emetogenic chemotherapy, and the all compared olanzapine combined with 5-HT3 RA plus dexamethasone compared with 5-HT3 RA plus dexamethasone for the prevention and treatment of CINV. Two different olanzapine dosage was used in the intervention group, six studies[19–21, 24, 26, 27]were 10 mg orally once a day, and four studies[12, 13, 22, 25] were 5mg oral once a day and Meng et al [23] was 2.5mg oral twice a day. Six studies[12, 13, 21–24] reported CINV according to the world health organization (WHO) classification standards for gastrointestinal reactions of anticancer drugs, and CINV □ (nausea without vomiting), CINV □ (vomiting 1-2 times a day, does not affect the eating), CINV □ (nausea and vomiting 3-5 times daily, affect eating, needed to treat), and CINV □ (uncontrollable nausea and vomiting, more than 5 times) were reported in articles. The main characteristics of the involved studies are presented in Table 1.

### Assessing Risk of Methodologic Quality

The detail of the risk-of-bias assessment was summarized in supplemental data 4. Except for Sandip 2017[19], all studies were open-label randomized study and assessed as high risk in allocation concealment. Four studies[12, 19, 20, 27] reported the detail of the randomized method, and the low risk was designated. Incomplete outcome data and selective reporting for all studies were assessed as low risk. In all, the overall methodologic quality was accepted.

### The Efficacy of Olanzapine for Prevention and Treatment of Delayed CINV

As shown in figure 1, eleven RCTs (1107 cancer patients) reported delayed CINV, and significant heterogeneity (*I*^2^=68%, *P*<0.1) was conducted, so a random model was used. There was significant lower rate of delayed CINV (RR=0.50, 95%CI: 0.38-0.66, *P*<0.01) when olanzapine was added in 5-HT3 RA and dexamethasone. Six articles (table 2) reported the level of delayed CINV, and statistically significant CINV □ (RR= 0.27, 95%CI: 0.14-0.52, *P*<0.01) and CINV □ (RR= 0.16, 95%CI: 0.04-0.60, *P*<0.01) were significantly decreased in olanzapine group. Subgroup analysis (table 3) shown that there was no statistical difference between 5mg and 10mg of olanzapine for delayed CINV.

**Figure 1.**
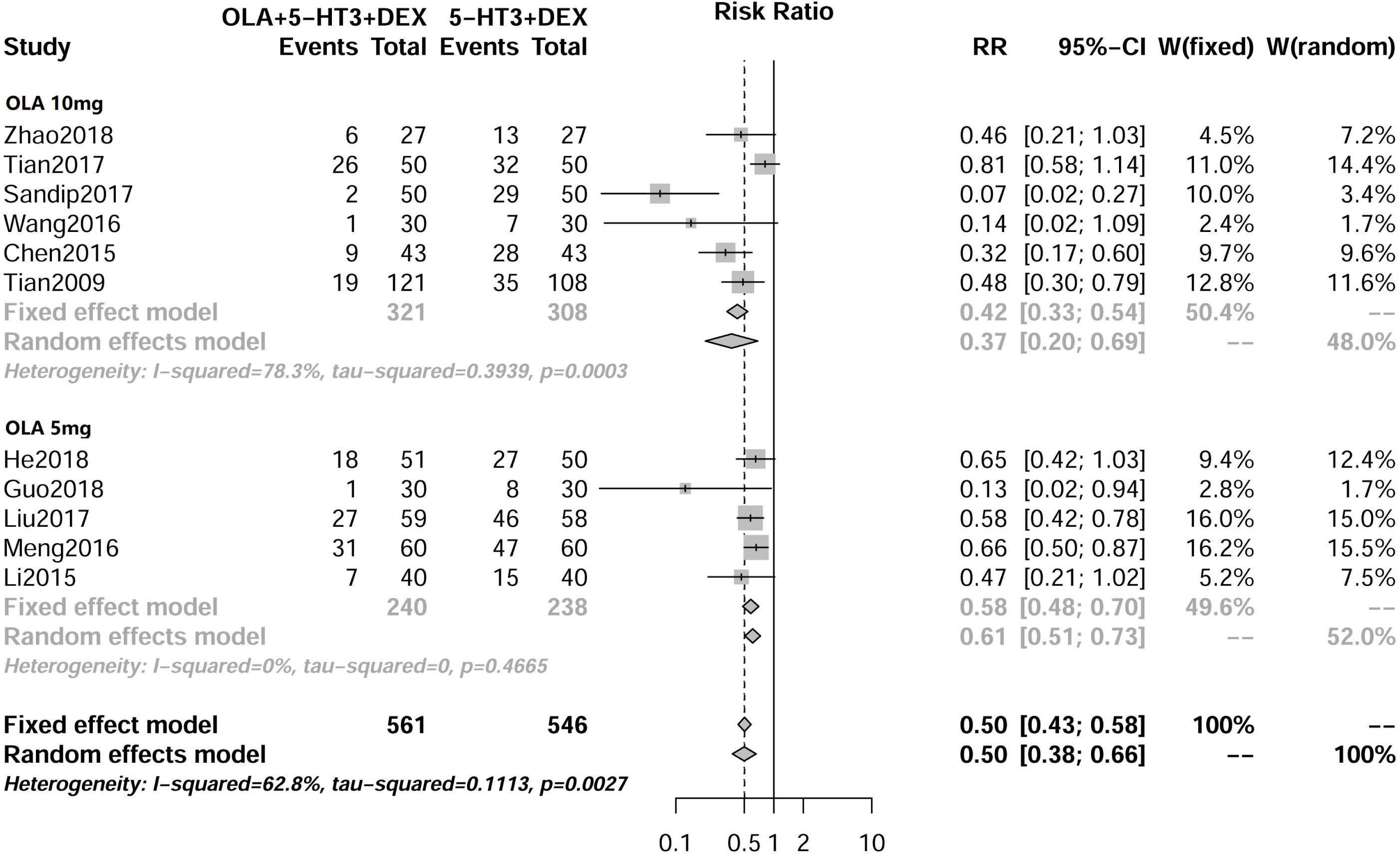
Meta-analysis result of the RR of delayed CINV. (RR= relative risk, CINV= chemotherapy-induced nausea, and vomiting).

### The AEs of Olanzapine for Prevention and Treatment of CINV

There were 7 AEs (dizzy, constipation, somnolence, anorexia, fatigue, insomnia and thirst) including in this systematic review to quantitative analysis, and anorexia and thirst found significant heterogeneity, so the random model was used. The rest of AEs did not detect significant heterogeneity, and a fixed model was used in this review. As shown in table 4, except for insomnia, the RR is 0.12 (95%CI: 0.06-0.26, *P*<0.01), all AEs were not conducted statistical difference between two groups. The results of subgroup analysis are following.

#### Dizzy

5 studies reported the dizzy events, no significant heterogeneity (*I*^2^=0.0%, *P*=0.97) was conducted, so fixed model was used. As showed in table 4, no significant difference was found between two groups (RR=1.14, 95%CI: 0.66-1.98, *P=*0.65), and the subgroup analysis shown that no statistical difference between olanzapine 10mg and 5mg (table 3).

#### Constipation

7 studies included in this review, and fixed model was used (*I*^2^=0.0%, *P*=0.25), and the rate of constipation was comparable (RR=1.13, 95%CI: 0.87-1.46, *P=*0.35) between two groups. No significant difference was detected by the subgroup analysis (table 3).

#### Anorexia

2 studies reported the data of anorexia, and significant heterogeneity (*I*^2^=68.1%, *P*=0.08) was found, so random model was used. The RR of anorexia was 1.73 (95%CI: 0.13-23.60, *P=*0.68), subgroup analysis was not conducted, because only one study in two groups.

#### Somnolence

4 studies reported the data of somnolence, and fixed model (*I*^2^=0.0%, *P*=0.61), and result showed that no significant difference was found (RR=1.20, 95%CI: 0.63-2.77, *P*=0.58), subgroup analysis did not found significant difference between 10mg and 5mg (table 3).

#### Fatigue

4 studies reported the fatigue events, no significant heterogeneity (*I*^2^=42.2%, *P*=0.85) was conducted, so fixed model was used. As showed in table 4, no significant difference was fund (RR=0.94, 95%CI: 0.61-1.43, *P*=0.76). All studies used 5mg olanzapine per day, so subgroup analysis was not conducted.

#### Insomnia

2 studies included in this review, and fixed model was used (*I*^2^=0.0%, *P*=0.45). As showed in table 4, the percentage of insomnia was significant lower in olanzapine group (RR=0.12, 95%CI: 0.06-0.26, *P*<0.01). Because of only two studies was involved, subgroup analysis was not conducted.

#### Thirst

2 studies reported the data of somnolence, and random model (*I*^2^=50.5%, *P*=0.16), and result showed that no significant difference was found (RR=1.24, 95%CI: 0.65-2.12, *P*=0.60). Because of only two studies was involved, subgroup analysis was not conducted.

#### The Efficacy of Olanzapine for Prevention and Treatment of Acute CINV

As shown in figure 2, 10 RCTs (990 cancer patients) provided the data of acute CINV, no statistically significant difference was conducted, and fixed model was used. The overall pooled RR of acute CINV was 0.60 (95%CI: 0.48–0.75, *P*<0.01). 6 articles (table 2) reported the level of acute CINV, while only CINV □ (RR= 0.26, 95%CI: 0.13-0.54, *P*<0.01) and CINV □ (RR= 0.17, 95%CI: 0.04-0.73, *P*=0.02) conducted statistically significant difference. Subgroup analysis (table 3) shown that no statistical difference between 5mg and 10mg of olanzapine for acute CINV was conducted.

**Figure 2.**
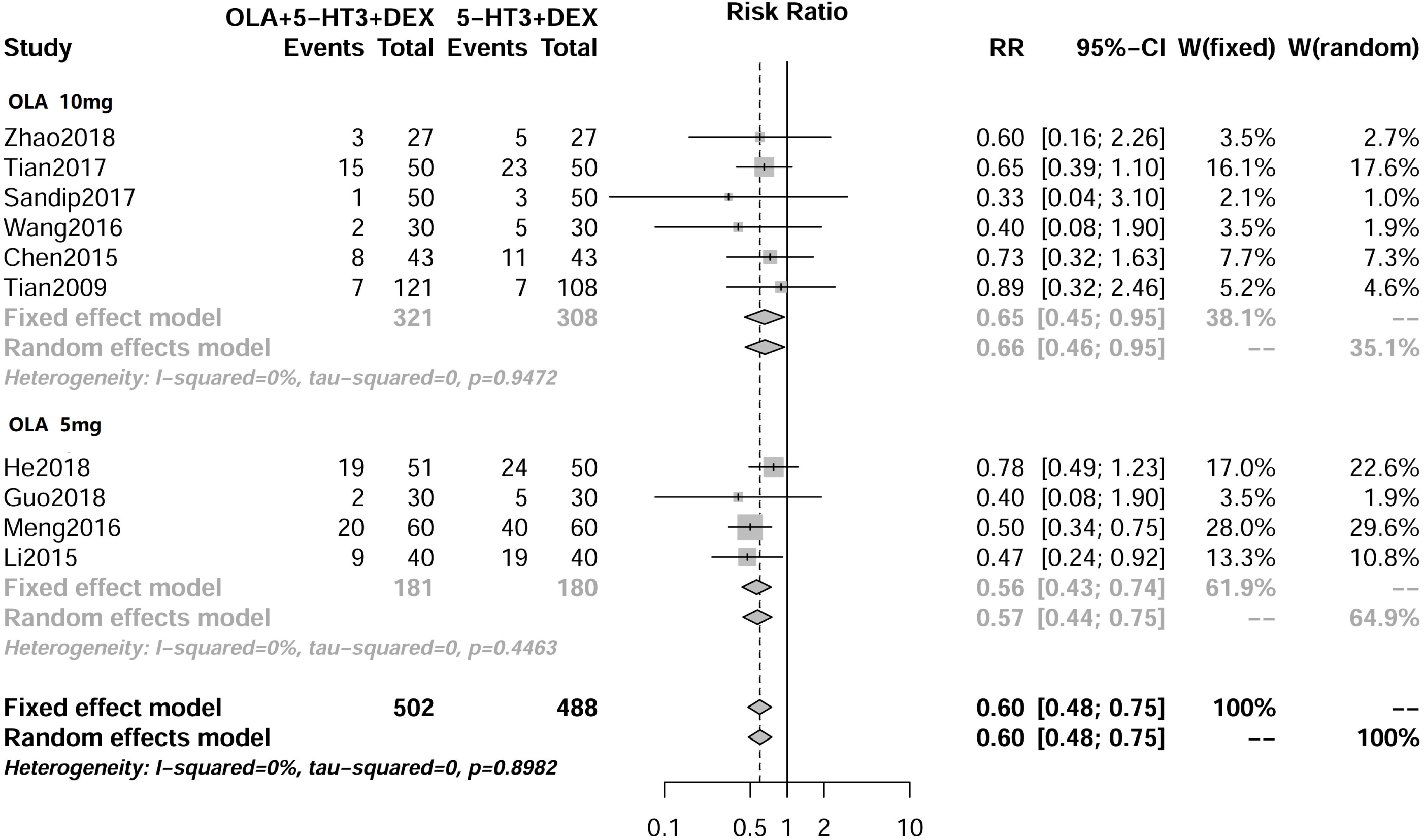
Meta-analysis result of the RR of acute CINV. (RR= relative risk, CINV= chemotherapy-induced nausea, and vomiting).

### Publication Bias

The publication bias of our meta-analysis was assessed using Beeg and Egger regression [28, 29], and evidence of publication bias was found from the statistical tests (Begg’s test, *P*=0.02, Egger’s test, *P*<0.01).

## Discussion

Eleven studies with 1107 cancer patients were involved in this review, and the result shows that olanzapine combined with 5-HT3 RA plus dexamethasone is more effect and less insomnia compared with 5-HT3 RA plus dexamethasone for the prevention and treatment of CINV in high and moderately emetogenic chemotherapy. Compared with 10mg per day, 5mg oral may be more appropriate for cancer patients.

CINV □ and CINV □ were greatly reduce the quality of life and need medical treatment. Six studies reported the level of CINV in our review, and the rate of CINV □ and CINV □ were significantly less in olanzapine groups. Besides olanzapine is still effective when aprepitant was failed in control CINV in patients receiving highly emetogenic chemotherapy [30], so olanzapine is an excellent alternative for the prophylaxis of CINV. The percentage of acute and delayed for olanzapine group was even lower than four drugs scheme[5] (acute CINV: 17.1% vs. 26.2%, delayed CINV: 26.2% vs. 57.6%), the possible reason may below: 1) our review involved high and moderately emetogenic chemotherapy, and Rudolph et al only include high emetogenic chemotherapy; 2) compared with Rudolph et al, the time of 5-HT3 RA and dexamethasone were longer (4-5 days vs. 1 day), and rate of CINV would be significant declined. More studies are warranted.

Two different olanzapine dosage was used in this review, and subgroup analysis shows that both doses of 10 mg and 5 mg olanzapine provided comparably significant improvement in acute and delayed CINV. The phase □ trial by Yanai et al[14]even find 5mg had a higher complete control in delayed CINV compared with 10mg (83.1% vs. 77.6%). Because olanzapine may lead to central nervous system depression, especially in elderly and debilitated patients[31], so 5 mg per day of olanzapine is more suitable for cancer patients.

Polled analysis is conducted in 7 AEs (dizzy, constipation, somnolence, anorexia, fatigue, insomnia and thirst) in this systematic review, and incidence of insomnia (RR=0.12, 95%CI: 0.06-0.26, *P*<0.01) is significantly lower in olanzapine groups. Insomnia is the most common AE of dexamethasone[32], olanzapine is an atypical antipsychotic agent that may lead drowsiness[33], so the patients with cancer had lower insomnia when olanzapine combined with NK-1 RA and dexamethasone. At the same time, olanzapine is not increased the incidence of somnolence, while no studies reported the severity and duration of AEs. Our further RCT about olanzapine combine tropisetron and dexamethasone for the prevention and treatment of nausea and vomiting induced by chemotherapy of patients with lung cancer (https://clinicaltrials.gov/ct2/show/NCT03571126?term=20180520&rank=1) concentrate on the level of AEs, and this concern may be solved. Subgroup analysis showed that there is no significant difference between 5mg and 10mg in dizzy, constipation and somnolence, so 5mg may be more appropriate. Previous phase 3 trial[5] found that there are no serious adverse events related to olanzapine, and no patient discontinued olanzapine because of toxic effects. Therefore olanzapine is safety in control CINV.

For the first time, we analyzed the level of CINV that is most concerned by patients and doctors, and the result shows that CINV III and CINV IV is significantly decreased when olanzapine were used. Besides, this review is less bias than previous studies [9, 10, 15, 34], because the intervention measure is more consistent in all involved articles, and we analyzed the efficacy and adverse AEs of olanzapine combined with 5-HT3 RA plus dexamethasone compared with 5-HT3 RA plus dexamethasone. Finally, our review involved the largest samples (1107 cancer patients) based on RCTs, so a higher level of evidence may be conducted. However, limitations also existed in this review. Not all studies reported the whole AEs, and the grade of AEs was not reported in included articles, the ongoing RCT conducted by our team will report relevant data. Besides, different types and dosage 5-HT3 RA were used in this review, and the efficacy and AEs was not all comparable between different type 5-HT3 RA[35], so potential bias may exsited. Finally, the use time of antiemetic drug and usage of dexamethasone were also not all consistent, which would affect the results in some degree.

In conclusion, olanzapine significant decreases the occurrence of CINV □ and □ and insomnia for the prevention and treatment of CINV in high and moderately emetogenic chemotherapy. Compared with 10mg per day, 5mg oral may be more appropriate for cancer patients. There is uncertainty about the severity and duration of AEs.

## Data Availability

All data of our review were extracted from the articles which published on journal.

## Disclosures and Acknowledgments

The authors have declared no conflicts of interest. Ethical approval and informed consent were not necessary because this is not primary research. This work was supported by the National Natural Science Foundation of China (Grant No.81360351, 81660512).

## Supporting Information

Table 1. Main characteristics of involved studies.

Table 2. Meta-analysis result of the RR of the level of acute and delayed CINV. (RR= relative risk, CINV= chemotherapy-induced nausea and vomiting).

Table 3. The results of subgroup analysis about olanzapine dosage.

Table 4. Meta-analysis result of the RR of AEs. (RR= relative risk, AEs=adverse events)

Supplemental Data 1. PRISMA Checklist.

Supplemental Data 2. The Search Strategy of PubMed.

Supplemental Data 3. Flow diagram of the details of the study.

Supplemental Data 4. Appraisal of risk of bias of the included trials using the Cochrane risk-of-bias tool. (Low risk=bias, if present, is unlikely to alter the results seriously, unclear risk=bias raises some doubt about the results, high risk=bias may alter the results seriously)

